# Transcutaneous auricular vagus nerve stimulation enhanced inhibitory control via increasing intrinsic prefrontal couplings

**DOI:** 10.1101/2023.09.14.23295532

**Authors:** Siyu Zhu, Qi Liu, Xiaolu Zhang, Menghan Zhou, Xinqi Zhou, Fangyuan Ding, Rong Zhang, Benjamin Becker, Keith M Kendrick, Weihua Zhao

## Abstract

Inhibitory control represents a core executive function that critically facilitates adaptive behavior and survival in an ever-changing environment. Non-invasive transcutaneous auricular vagus nerve stimulation (taVNS) has been hypothesized to improve behavioral inhibition performance, however the neurocomputational mechanism of taVNS-induced neuroenhancement remain elusive. In the current study, we investigated the effect of taVNS on inhibitory control in a pre-registered sham-controlled between-subject functional near infrared spectroscopy (fNIRS) experiment with an emotional face Go/No-Go paradigm in ninety subjects. After data quality check, eighty-two subjects were included in the final data analysis. Behaviorally, the taVNS improved No-Go response accuracy, together with computational modeling using Hierarchical Bayesian estimation of the Drift Diffusion Model (HDDM) indicating that it specifically reduced the information accumulation rate for Go responses, and this was negatively associated with increased accuracy of No-Go responses. On the neural level, taVNS enhanced engagement of the bilateral inferior frontal gyrus (IFG) during inhibition of angry expression faces and modulated functional couplings (FCs) within the prefrontal inhibitory control network. Mediation models revealed that taVNS-induced facilitation of inhibitory control was critically mediated by a decreased information accumulation for Go responses and concomitantly enhanced neurofunctional coupling between the inferior and orbital frontal cortex. Our findings demonstrate a striking potential for taVNS to improve inhibitory control via reducing pre-potent responses and enhancing FCs within prefrontal inhibitory control networks, suggesting a promising therapeutic role in treating specific disorders characterized by inhibitory control deficits.

## 1. Introduction

Inhibitory control is a core executive function vital for adaptive behavioral regulation via suppression of inappropriate responses. In everyday life it allows us to control automatic urges at perceptual, cognitive, and behavioral levels (Diamond, 2013). Prefrontal cortical (PFC) circuits critically implement inhibitory control on the neural level (Goldstein *et al*., 2007), particularly the inferior frontal gyrus (IFG) (Munakata *et al*., 2011; Zhuang *et al*., 2022). Interestingly, deficits in inhibitory control (e.g. impulsivity, hyperactivity) are the primary transdiagnostic characteristics of individuals with attention-deficit/hyperactivity disorder (ADHD) (Polanczyk *et al*., 2007), substance use disorders (Hildebrandt *et al*., 2021), posttraumatic stress disorder (PTSD) (Catarino *et al*., 2015) and obesity (Jasinska *et al*., 2012). Improving inhibitory control thus represents a highly promising therapeutic target for clinical application.

Transcutaneous auricular vagus nerve stimulation (taVNS) - a novel non-invasive neuromodulation technique - has been hypothesized to promote inhibitory control via its regulation of the locus coeruleus-norepinephrine (LC-NE) network and GABAergic system (Burger *et al*., 2020; Zhu *et al*., 2022b). The LC-NE network plays a pivotal role in inhibitory control. Accumulating evidence from brain imaging studies indicates that the neural activity of LC-NE system could modulate functional connectivity within the prefrontal inhibitory control network (Chamberlain *et al*., 2007, 2009; Passamonti *et al*., 2018; Li *et al*., 2020; Tomassini *et al*., 2022). Recently, one study reported that oral atomoxetine (i.e. a noradrenergic reuptake inhibitor) improved reaction times during inhibition in Parkinson patients with lower LC integrity (O’Callaghan *et al*., 2021). Consistent with this, taVNS has been found to increase the activation of brainstem regions, including the LC (Frangos *et al*., 2015; Yakunina *et al*., 2017), suggesting a modulatory role of taVNS in inhibitory control ability via its impact on the LC-NE network. Moreover, the neurotransmitter γ-amnobutyric acid (GABA) also plays a key role in modulating cognitive performance with demands for inhibitory control. Using magnetic resonance spectroscopy, Hermans and colleagues found that older adults with lower GABA levels exhibited a prolonged stop-signal responses time (Hermans *et al*., 2018), indicating a negative relationship between inhibitory response and GABA levels, particularly in inferior frontal regions (Murley *et al*., 2020). More importantly, taVNS significantly increased GABA-A receptor activity (Capone *et al*., 2015). Taken together, these findings suggest that taVNS may be an effective neuromodulator to improve inhibitory control by regulating activity of the LC-NE network and GABAergic system.

Some initial studies provided preliminary although inconsistent evidence for beneficial effects of taVNS on inhibitory control. For instance, although Borges et al (2020) suggested that taVNS can increase cognitive flexibility in a set-shifting task, no improvement on inhibitory performance in either the Flanker task (i.e. for selective attention measurement) or in the Spatial Stroop task were found (Borges *et al*., 2020). However, it has been subsequently reported that taVNS improved adaption to conflict in the Simon task (Fischer *et al*., 2018). Additionally, although some initial behavioral studies have consistently shown that response inhibition in a Go/No-Go task was enhanced by active taVNS (Beste *et al*., 2016; Keute *et al*., 2020; Pihlaja *et al*., 2020), the neural mechanism of the potential beneficial effects of taVNS on inhibition has not been explored.

To better elucidate the potential for taVNS to enhance inhibitory control performance and the underlying neurocomputational mechanism, we here investigated the impact of taVNS on inhibition ability in combination with functional near-infrared spectroscopy (fNIRS). fNIRS is increasingly used as an optical neuroimaging method based on the hemodynamic response (i.e. concentration of oxygenated hemoglobin; HbO) due to a higher temporal resolution and lower sensitivity to movement artifacts which are important for fast response tasks such as the Go/No-Go task (Ferrari and Quaresima, 2012; Sakai, 2022). In the present study, we adopted a modified emotional Go/No-Go paradigm with neutral expression faces as Go and emotional expression faces (i.e., angry and happy faces) as No-Go stimuli. Most importantly, we further applied a well-validated computational model, the diffusion decision model (DDM), which characterizes within- and between-subject differences in the Go/No-Go paradigm (Gomez *et al*., 2007; Zhang *et al*., 2016; Huang-Pollock *et al*., 2017; Ratcliff *et al*., 2018; de Gee *et al*., 2020; Weigard *et al*., 2020; Gorka *et al*., 2022) to fully reveal the critical role of taVNS in modulating inhibitory control performance from both neurocomputational and behavioral levels. Overall, we hypothesized that taVNS relative to a sham-control stimulation (earlobe) would enhance behavioral inhibition in the emotional Go/No-Go task and that this would be associated with altered neural responses and connectivities in the prefrontal cortex circuitry important for executive control.

## 2. Materials and methods

### 2.1. Participants

We recruited 90 healthy adult Chinese university students for the current study with all reporting being free from medical or psychiatric disorders or current or regular medication, and who were required not to consume any alcohol, caffeine or nicotine on the day of the experiment. All participants had normal or corrected to normal vision. Data from 8 participants were excluded due to not recognizing facial emotions in the Go/No-Go task (n=5) or technical problems of recording fNIRS data (n=3), leaving a total of 82 participants (42 females, mean age 19.61±2.01 years) for the final analyses. Each participant provided written informed consent for the study protocol approved by the ethical committee of the University of Electronic Science and Technology of China. The study was pre-registered as a clinical trial (ClinicalTrials.gov ID: NCT05468385).

### 2.2. Procedure

In a sham-controlled, single-blind, between-subject design, participants were randomly assigned to two groups, receiving real taVNS or sham stimulation respectively. Upon arrival, all subjects completed a number of validated psychometric questionnaires (details see in supplementary information) to exclude confounding effects of personality traits. In addition the Positive and Negative Affect Schedule(PANAS) was administered twice (before and immediately after the experiment) to access mood changes (Watson *et al*., 1988). Next, stimulation intensity was adjusted according to participants’ subjective feelings (see section 2.3). Participants were then asked to rest for 5 minutes after familiarizing themselves with the emotional Go/No-Go tasks. Subsequently, resting-state brain activity was recorded using fNIRS while participants were instructed to relax and fixate on a white cross centered on the black screen during 15-minute period of stimulation (not reported here). Finally, following another 15 minutes’ stimulation, participants were asked to complete the emotional Go/No-Go task (see section 2.4). At the end of the experiment, subjects were asked to report side effects of stimulation including headache, nausea, skin irritation under the electrode, relaxed, vigilant, unpleasant feelings, dizziness, neck pain, muscle contractions in the neck, and stinging sensation in the ear on a seven-point Likert scale (1: not at all, to 7: very much). An illustration of the procedure is presented in **Fig.1A**.

**Fig. 1.**
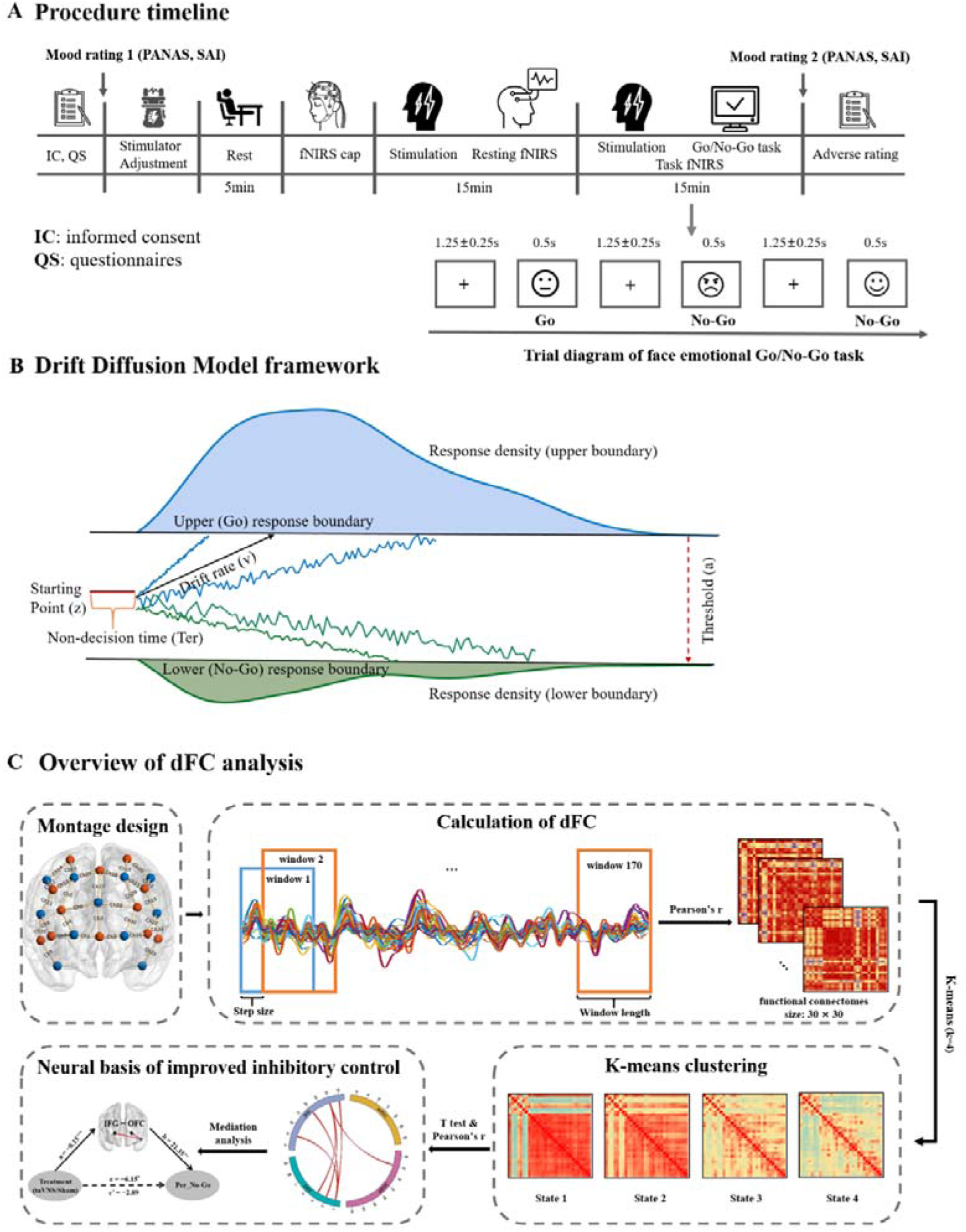
Schematic illustration of experimental protocol and data analysis. A, Procedure timeline. B, Drift Diffusion Model framework. C, Overview of dynamic functional connectivity (dFC) analysis.

### 2.3. Transcutaneous auricular vagus nerve stimulation

In line with our previous study (Zhu *et al*., 2022a), taVNS was implemented via a modified transcutaneous electrical acupoint stimulation device with an ear-clip electrode being attached to the left tragus for vagal stimulation in the taVNS group and to the left earlobe for sham stimulation in the control group. Electrical pulses (width, 500 μs; frequency, 25 Hz) were delivered for 30 s, alternated by a 30 s pause for a total duration of 30 minutes (i.e., two stimulation periods with each lasting 15 minutes). The auricular branch of vagus nerve (VN) is related to touch sensation, stimulation intensity was therefore individually calibrated to a level above the detection threshold but not generating any discomfort feeling to ensure VN activation (Ellrich, 2011). During the calibration procedure, participants received increasing and decreasing series of stimulation trials, and reported their subjective sensation of the stimulation on a 7-point Likert scale (1: feeling nothing, to 7: painful). The increasing series of trials started from 0 mA and elevated in steps of 0.1 mA until participants reported a “painful” sensation of 7. On the other hand, in decreasing series of trials, the “painful” intensity was repeated and then reduced in steps of 0.1 mA until a subjective sensation of “feeling nothing” was experienced. This procedure was performed twice and the final stimulation intensity for each participant was calculated based on the average of four intensity values (i.e., two from increasing and two from decreasing trials) that were rated as 5 (i.e., obvious “tingling” sensation but not painful). The average stimulation intensity is 0.77 mA (0.3-1.2 mA) for the taVNS group, and is 0.94 mA (0.6-1.4 mA) for the sham-controlled group.

### 2.4. Emotional Go/No-Go task

The present study adopted an emotional face Go/No-Go task (**see Fig. 1A**). Each trial consisted of a white fixation cross (1250 ± 250 ms) on a black background and a random presentation of emotional faces (i.e., angry, happy and neutral) for 500 ms in the center of a 24-inch monitor at a resolution of 1024×768 pixels (60Hz) using E-prime 3.0 (Psychology Software Tools, Inc). Participants were required to respond to neutral faces as “Go” stimuli by pressing the button as fast as possible (i.e., before next trial) but to withhold responses when angry or happy faces as “No-Go” stimuli were presented. All 96 face images (48 neutral, 24 angry and 24 happy) were grayscale images with equivalent size and cumulative brightness, which were selected from the Chinese facial affective picture system (Gong *et al*., 2011). This task consisted of 144 “Go” trials (each neutral face was presented 3 times) and 48 “No-Go” trials (each angry or happy face was presented once).

### 2.5. Measurements

#### 2.5.1 Behavioral performance and computational modeling of inhibitory ability

To evaluate the effects of taVNS on response inhibition, behavioral performance including reaction time in Go trials (RT_Go) and accuracy of No-Go trials (ACC_No-Go) were measured. We also explored inhibitory ability using Hierarchical Bayesian estimation of the Drift Diffusion Model (HDDM, implemented in Python 3.8 (Wiecki *et al*., 2013)). The HDDM was fit to trial-by-trial measures of response type (i.e., “Go” vs. “No-Go”) and reaction times, and four parameters including drift rate (v, the rate of information accumulation toward the correct choice), starting point (z, a response bias for “Go” response or “No-Go” response), boundary separation (a, how much information is required to make a decision (i.e. the separation between the upper and lower boundary), and non-decision time (Ter, which reflects aspects of processing unrelated to decision making and represents sensory encoding and motor execution) were estimated across taVNS and sham-controlled groups respectively. For details of model framework **see Fig. 1B**, model estimation and model simulation see in Supplementary information.

#### 2.5.2 fNIRS data collection and neural measurements

During the emotional Go/No-Go task, a NIRSport2 system (NIRx Medical Technologies LLC, Berlin, Germany) was utilized to measure the hemodynamic activity of each participant at a sampling frequency of 6.78 Hz. Thirty channels (12 sources and 11 detectors) were placed bilaterally over the prefrontal cortex (**Fig. 1C**) based on fNIRS Optodes’ location decider toolbox (fOLD v2.2) (Zimeo Morais *et al*., 2018). Notably, these regions are highly related to emotional inhibitory control based on previous studies (Munakata *et al*., 2011; Zhuang *et al*., 2021), including orbitofrontal cortex (OFC), inferior frontal gyrus (IFG), medial prefrontal cortex (mPFC), and dorsolateral prefrontal cortex (dlPFC). Each source-detector pair defined a single measurement channel with a distance of 3.0 cm and placement of fNIRS optodes was according to the 10-10 International System. Hair was manually parted under the optodes to improve signal detection.

### 2.6. Data analyses

#### 2.6.1 Self-reported measures

To exclude the potential confounding effects of self-reported mood and personality traits, independent *t* tests were performed separately with treatment (taVNS vs. sham) as the between group variable. Independent *t* tests for the side effect ratings between taVNS and sham stimulation group were also performed.

#### 2.6.2 Behavioral performance and computational model indices analyses

Firstly, independent *t* tests were used to compare taVNS and sham groups for model-free indices (i.e., RT_Go, and ACC_No-Go) and HDDM-based indices (a, v, z, and Ter). Pearson correlation analyses were performed to investigate the relationship between the accuracy of No-Go and model-based indices. Finally, we further conducted a mediation analysis to investigate whether the drift rate for Go stimuli mediated the effect of taVNS on response inhibition performance by means of a bootstrapping method.

#### 2.6.3 Functional NIRs data analyses

The fNIRS data were preprocessed and converted to time series of oxyhemoglobin (HbO) for each channel using NIRS-KIT (Hou *et al*., 2021) (Supplementary Methods: fNIRS data preprocessing). Given HbO is a more sensitive indicator of task-associated changes relative to deoxyhemoglobin (HbR) (Ferrari and Quaresima, 2012), we only focused on HbO in the further analyses.

##### 2.6.3.1 Generalized linear model analyses

The contrasts between “No-Go” and “Go” trials (i.e., angry vs. neutral, and happy vs. neutral) within 7 ROIs (i.e., left & right OFC, left & right IFG, left & right dlPFC and mPFC) were measured at the individual-level using generalized linear model (GLM) approach (see Supplementary information). The data were then subjected to a group-level analysis by means of repeated-measures ANOVA with ROI and emotion as two within-subject factors, and treatment as a between-subject factor. Bonferroni correction was applied to post hoc comparison tests.

##### 2.6.3.2 fNIRS-Based dynamic functional connectivity analyses

Dynamic functional connectivity was constructed using a sliding-window correlation analysis, and k-means clustering was applied to generate the key brain connectivity states under taVNS and sham conditions, respectively (Tang *et al*., 2021; Lu *et al*., 2023). Firstly, dynamic functional connectivity analysis using the sliding window method (window length: 252 data points, ∼37.1s) was conducted based on time series of HbO data from a total of 30 channels per participant to investigate the transient functional coupling (FC) within prefrontal regions during the whole emotional Go/No-Go task. Further, we adopted k-means algorithm (L1 distance) to cluster all the calculated functional connectomes to estimate the reoccurring functional states (patterns) of task-based time-varying oscillations of HbO signal (details see Supplementary information). Finally, treatment groups were compared on FCs within each state using independent *t* tests with permutations for multiple comparisons (Nichols and Holmes, 2002; Camargo *et al*., 2008). We further employed Pearson correlation with permutation tests (10000 permutations) to identify which prefrontal FCs was highly contributed to inhibition performance. Mediation analysis was performed to investigate whether these FCs mediated taVNS effects on inhibition ability. Overview of the analyses was illustrated in **Fig. 1C**.

## 3. Results

### 3.1. Comparable participant characteristics in the treatment groups

Participant characteristics are summarized in **Table 1**, the two treatment groups did not differ significantly in terms of age (*p*= 0.744), gender (*p* =0.659), mood (*p*s >0.15), personality traits (*p*s >0.17), and subjective ratings for the stimulation adverse effects (*p*s >0.13).

**Table 1.**
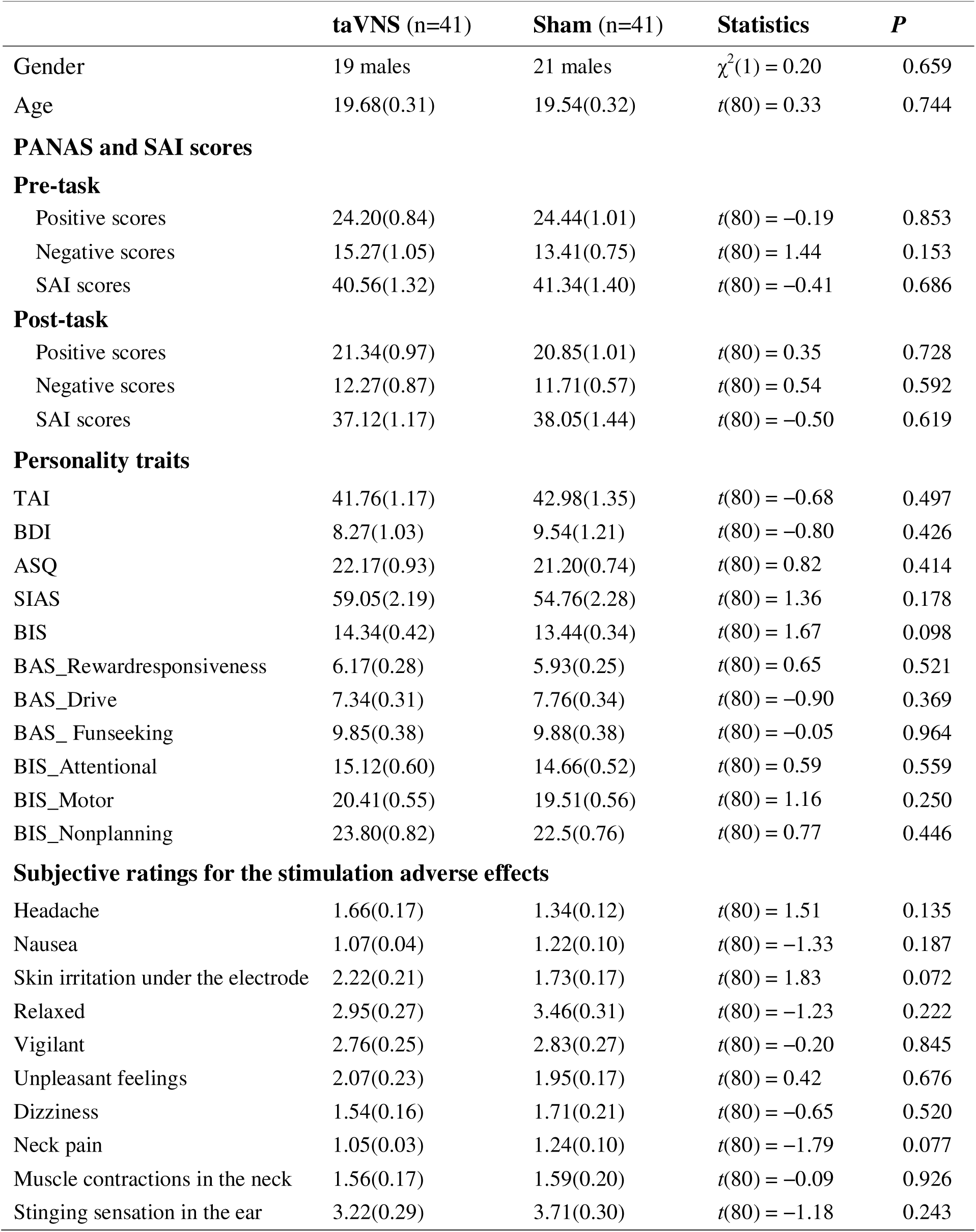
Sample characteristics and self-reported measurements (mean ± SEM).

|  | taVNS (n=41) | Sham (n=41) | Statistics | *P* |
| --- | --- | --- | --- | --- |
| Gender | 19 males | 21 males | $\chi^2(1) = 0.20$ | 0.659 |
| Age | 19.68(0.31) | 19.54(0.32) | $t(80) = 0.33$ | 0.744 |
| **PANAS and SAI scores** |  |  |  |  |
| **Pre-task** |  |  |  |  |
| Positive scores | 24.20(0.84) | 24.44(1.01) | $t(80) = -0.19$ | 0.853 |
| Negative scores | 15.27(1.05) | 13.41(0.75) | $t(80) = 1.44$ | 0.153 |
| SAI scores | 40.56(1.32) | 41.34(1.40) | $t(80) = -0.41$ | 0.686 |
| **Post-task** |  |  |  |  |
| Positive scores | 21.34(0.97) | 20.85(1.01) | $t(80) = 0.35$ | 0.728 |
| Negative scores | 12.27(0.87) | 11.71(0.57) | $t(80) = 0.54$ | 0.592 |
| SAI scores | 37.12(1.17) | 38.05(1.44) | $t(80) = -0.50$ | 0.619 |
| **Personality traits** |  |  |  |  |
| TAI | 41.76(1.17) | 42.98(1.35) | $t(80) = -0.68$ | 0.497 |
| BDI | 8.27(1.03) | 9.54(1.21) | $t(80) = -0.80$ | 0.426 |
| ASQ | 22.17(0.93) | 21.20(0.74) | $t(80) = 0.82$ | 0.414 |
| SIAS | 59.05(2.19) | 54.76(2.28) | $t(80) = 1.36$ | 0.178 |
| BIS | 14.34(0.42) | 13.44(0.34) | $t(80) = 1.67$ | 0.098 |
| BAS_Rewardresponsiveness | 6.17(0.28) | 5.93(0.25) | $t(80) = 0.65$ | 0.521 |
| BAS_Drive | 7.34(0.31) | 7.76(0.34) | $t(80) = -0.90$ | 0.369 |
| BAS_Funseeking | 9.85(0.38) | 9.88(0.38) | $t(80) = -0.05$ | 0.964 |
| BIS_Attentional | 15.12(0.60) | 14.66(0.52) | $t(80) = 0.59$ | 0.559 |
| BIS_Motor | 20.41(0.55) | 19.51(0.56) | $t(80) = 1.16$ | 0.250 |
| BIS_Nonplanning | 23.80(0.82) | 22.5(0.76) | $t(80) = 0.77$ | 0.446 |
| **Subjective ratings for the stimulation adverse effects** |  |  |  |  |
| Headache | 1.66(0.17) | 1.34(0.12) | $t(80) = 1.51$ | 0.135 |
| Nausea | 1.07(0.04) | 1.22(0.10) | $t(80) = -1.33$ | 0.187 |
| Skin irritation under the electrode | 2.22(0.21) | 1.73(0.17) | $t(80) = 1.83$ | 0.072 |
| Relaxed | 2.95(0.27) | 3.46(0.31) | $t(80) = -1.23$ | 0.222 |
| Vigilant | 2.76(0.25) | 2.83(0.27) | $t(80) = -0.20$ | 0.845 |
| Unpleasant feelings | 2.07(0.23) | 1.95(0.17) | $t(80) = 0.42$ | 0.676 |
| Dizziness | 1.54(0.16) | 1.71(0.21) | $t(80) = -0.65$ | 0.520 |
| Neck pain | 1.05(0.03) | 1.24(0.10) | $t(80) = -1.79$ | 0.077 |
| Muscle contractions in the neck | 1.56(0.17) | 1.59(0.20) | $t(80) = -0.09$ | 0.926 |
| Stinging sensation in the ear | 3.22(0.29) | 3.71(0.30) | $t(80) = -1.18$ | 0.243 |

### 3.2. Effects of taVNS on response inhibition performance and its computational mechanism

Repeated measures ANOVA was conducted for ACC_No-Go with emotion (i.e., angry and happy) as a within-subject factor and treatment as group factor, independent *t* tests were performed to investigate taVNS effects on RT_Go and HDDM-based indices (a, v, z, and Ter) respectively. Results showed no significant interaction between emotion and treatment (*F*_[1, 80]_ = 5.12, *p* = 0.256) but a significant treatment effect on ACC_No-Go (*F*_[1, 80]_ = 5.18, *p* = 0.026, partial ☐*^2^* = 0.061), indicating an increased accuracy of No-Go trials following taVNS compared to sham treatment. The ANOVA results did not change with the stimulation intensity as a covariate factor. No change in reaction time for correct Go trials was found under treatment (*t*_[80]_ = 0.38, *p* = 0.703, **Fig. 2A**). Additionally, drift rate for Go trials decreased in the taVNS relative to the sham group (*t*_[80]_ = −2.29, *p* = 0.025, Cohen’s *d* = 0.506, **Fig. 2B**) and there were no group differences in the rest of the HDDM indices (a: *t*_[80]_ = −0.50, *p* = 0.621; v_nogo: *t*_[80]_ = −0.60, *p* = 0.552; Ter: *t*_[80]_ = 0.50, *p* = 0.618; z: *t*_[80]_ = 1.26, *p* = 0.212, each individual plot of observed vs. simulated responses across taVNS and sham is shown in Supplementary information **Figure S1-S18**), and an example of the HDDM simulation results are presented in **Fig. 3**. Mediation results showed that taVNS decreased drift rate for Go stimuli (path *a* = 0.57, *p* = 0.025) and drift rate was negatively associated with accuracy of No-Go trials (path *b* = −2.95, *p* = 0.014). Importantly, the indirect effect (path a×b) reached significance, suggesting drift rate for Go stimuli mediated taVNS effects on increasing inhibition performance (indirect effect = −1.68, 95% CI = [−3.98, −0.10], bootstrap = 5000, see **Fig. 2C**).

**Fig. 2.**
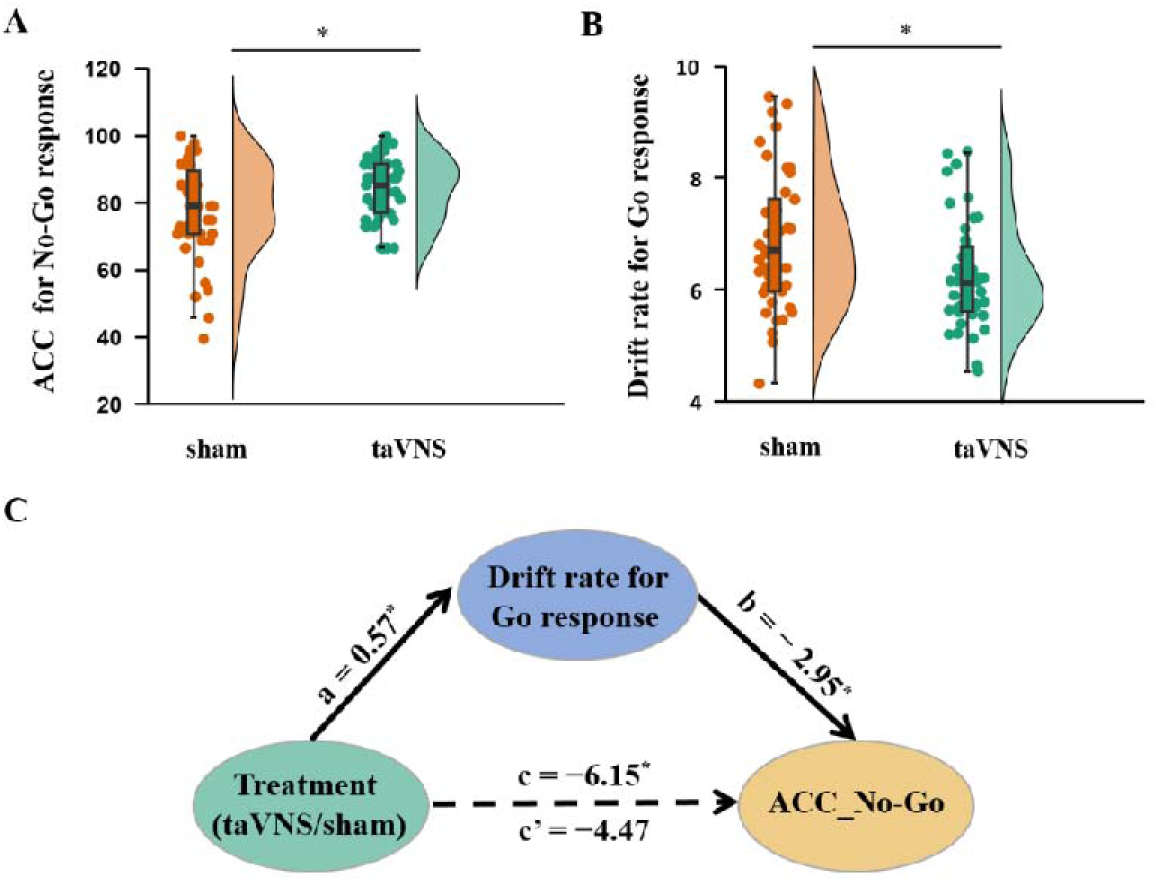
taVNS effects on behavioral index. A, Accuracy of No-Go response (mean ± SEM) under two treatment groups. B, Computed Hierarchical Bayesian estimation of the Drift Diffusion Model (HDDM) parameter differences between treatment groups. C, Drift rate of Go response mediated taVNS effects on the accuracy of No-Go response. \**p* < .05. \*\**p* < .01, \*\*\**p* < .001, ns: no significant difference.

**Fig. 3.**
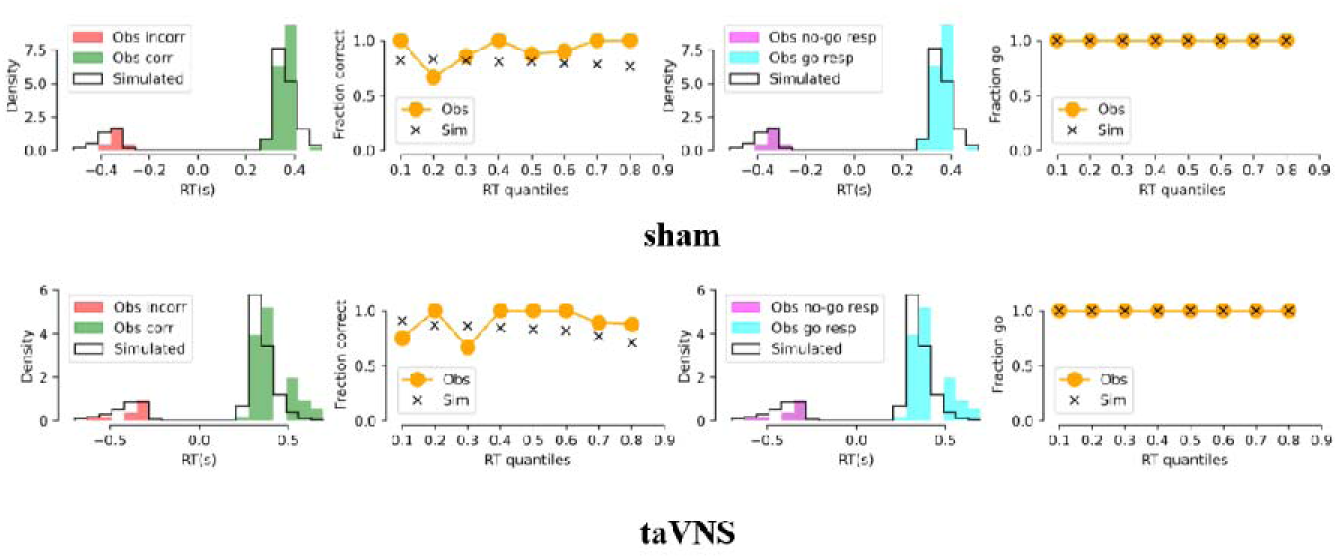
HDDM simulation results. Representative plot of observed vs. simulated responses (i.e., accuracy and reaction times) across taVNS and sham condition. Columns 1 and 3 represent probability densities. Columns 2 and 4 represent percentage of correct responses in each reaction time quantile. Obs = observed. Sim = simulated. Corr = correct. Incorr = incorrect.

### 3.3. taVNS effects on brain activity during response inhibition

A three-way repeated-measures ANOVA was performed on brain activity with ROI and emotion (i.e., angry vs. neutral, happy vs. neutral) as two within-subject factors, and treatment as a between-subject factor. Results revealed there was a significant three-way interaction effect (*F*_[6, 480]_ = 2.69, *p* = 0.033, partial ☐*^2^* = 0.033), suggesting that taVNS increased the activity of both right IFG (*p* = 0.015, Cohen’s *d* = 0.547) and left IFG (*p* = 0.044, Cohen’s *d* = 0.453, Bonferroni correction) in response to angry No-Go faces (**Fig. 4A**). In addition, the results did not change if we averaged the left and right regions (i.e. 4 ROIs, IFG, mPFC, dlPFC and OFC, *F*_[3, 240]_ = 3.271, *p* = 0.031, partial ☐*^2^* = 0.039).

**Fig. 4.**
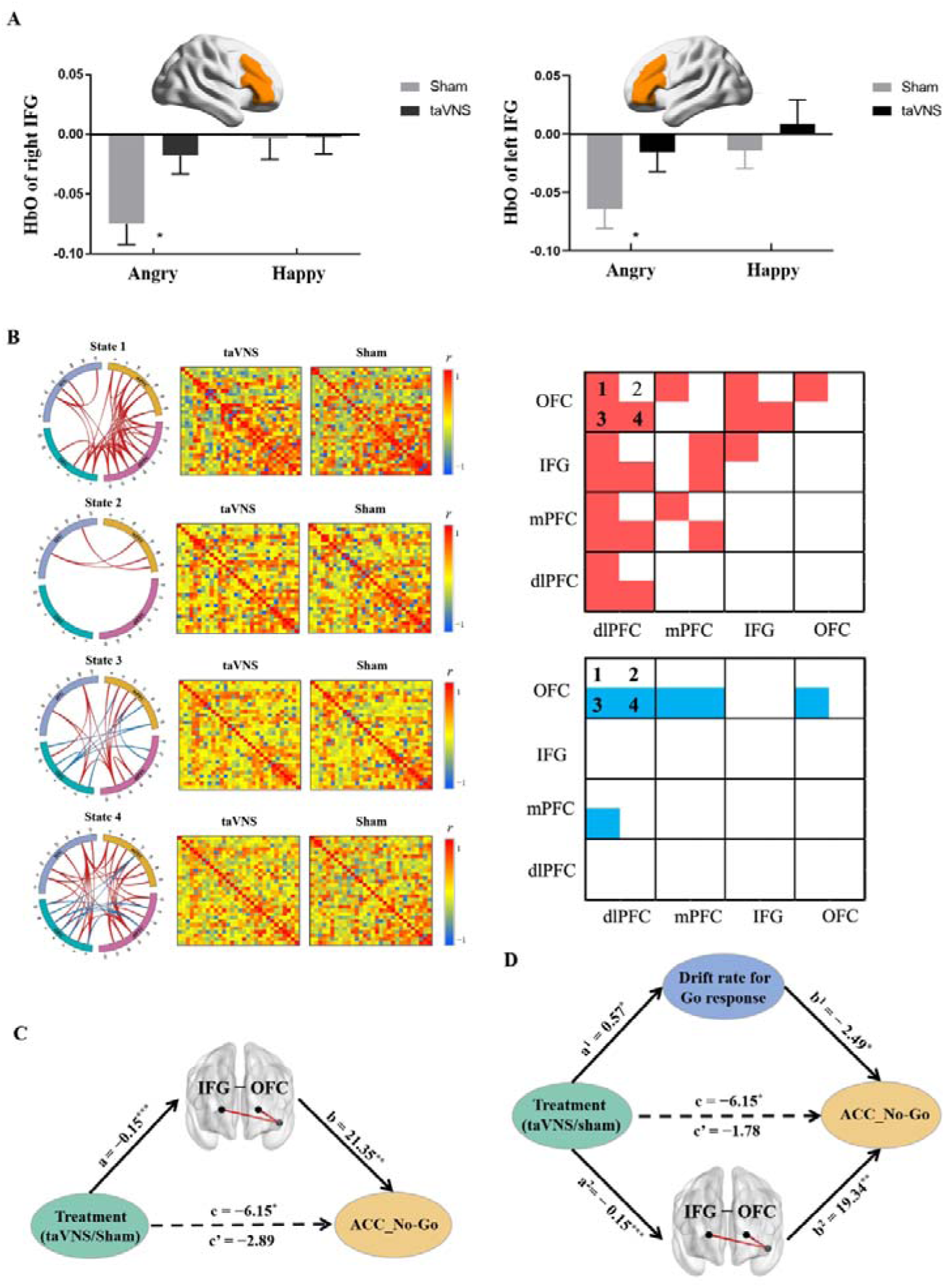
fNIRS results. A, HbO differences of bilateral IFG for inhibition on angry and happy faces under taVNS and sham treatment. B, Different effects of treatment on FCs in determined State 1, State 2, State 3, and State 4 respectively, red line/box: taVNS > sham; blue line/box: taVNS < sham. C, Mediation analysis between treatment, FC between IFG and OFC, and accuracy of No-Go response. D, Parallel mediation analysis between treatment, FC between IFG and OFC, drift rate for Go stimuli and accuracy of No-Go response. FC-functional connectivity; IFG-Inferior frontal gyrus; OFC-Orbitofrontal cortex. \**p* < .05, \*\**p* < .01, \*\*\**p* < .001.

### 3.4. taVNS effects on task-related prefrontal functional couplings

Four states were determined by k-means clustering and independent permutation *t* test (10000 permutations) results showed that taVNS increased both between FCs including dlPFC-IFG/OFC/mPFC, OFC-IFG/mPFC couplings and within FCs among these four regions in state 1, and increased IFG-mPFC FCs in state 2. Notably, taVNS increased FCs including IFG-OFC/dlPFC, dlPFC-mPFC/OFC and FC within dlPFC while decreasing specific FCs including mPFC-dlPFC/OFC, OFC-dlPFC, and FC within OFC in state 3. In state 4, taVNS increased FCs including IFG -OFC/dlPFC/mPFC, dlPFC-OFC/mPFC, and FCs within dlPFC and mPFC while decreasing specific FCs including OFC-dlPFC/mPFC (all *p*s< 0.05, **Fig. 4B** and more details see Supplementary information Table S1-Table S4).

Mediation analyses indicated that taVNS increased FC between IFG and OFC in state 4 (path *a* = −0.15, *p* < 0.001) and this FC was positively correlated with increased ACC_No-Go (path *b* = 21.35, *p* = 0.002). Notably, the indirect effect was significant showing that the coupling between IFG and OFC in state 4 mediated taVNS effects on increasing responses inhibition performance (indirect effect = −3.26, 95% CI = [−6.92, −0.61], bootstrap = 5000, see **Fig. 4C**).

### 3.5. Computational modeling of taVNS effects on inhibition performance

To further determine the underlying neurocomputational mechanism of taVNS-induced enhanced response inhibition, a parallel mediation analysis was conducted with drift rate for Go stimuli and FC between IFG and OFC in state 4 serving as mediating variables. The results revealed that the total effect of treatment on accuracy of No-Go trials was significant (path *c* = −6.15, *p* = 0.026) while the direct effect was not (path *c* ’= −1.78, *p* = 0.518). Specifically, the impact of taVNS on inhibition performance was mediated by drift rate for Go stimuli (indirect effect 1 = −1.42, 95% CI = [−3.51, −0.02]) and FC between IFG and OFC in state 4 (indirect effect 2 = −2.95, 95% CI = [−6.53, −0.32). A further comparison regarding the size of the mediation effect (Indirect 1 − Indirect 2 =1.53, 95% CI = [−1.67, 5.24], bootstrap = 5000) showed that decreased drift rate for Go stimuli and enhanced FC between IFG and OFC equally mediated the effect of taVNS on response inhibition (see **Fig. 4D**).

**Fig. 4** fNIRS results. A, HbO differences of bilateral IFG for inhibition on angry and happy faces under taVNS and sham treatment. B, Different effects of treatment on FCs in determined State 1, State 2, State 3, and State 4 respectively, red line/box: taVNS > sham; blue line/box: taVNS < sham. C, Mediation analysis between treatment, FC between IFG and OFC, and accuracy of No-Go response. D, Parallel mediation analysis between treatment, FC between IFG and OFC, drift rate for Go stimuli and accuracy of No-Go response. FC-functional connectivity; IFG-Inferior frontal gyrus; OFC-Orbitofrontal cortex. \**p* < .05, \*\**p* < .01, \*\*\**p* < .001.

## 4. Discussion

In the current study, we explored the potential of taVNS as a non-invasive neuromodulation technique to enhance inhibitory control. In line with our hypothesis, taVNS improved inhibition accuracy and increased the activation of bilateral IFG for inhibition during presentation of angry expression faces. During the entire emotional Go/No-Go task, four states were determined by k-means clustering and taVNS modulated the functional coupling within four key regions including OFC, dlPFC, mPFC and IFG regions in the respective states. Most importantly, mediation results suggested that the decreased rate of information accumulation for correct Go response calculated by HDDM computational modeling combined with increased IFG-OFC functional coupling equally mediated the taVNS-induced enhancement of inhibition.

In line with preliminary evidence (Beste *et al*., 2016; Keute *et al*., 2020; Pihlaja *et al*., 2020), our study found that taVNS increased accuracy of correct No-Go responses (i.e., ACC_No-Go) compared to sham stimulation, indicating a beneficial effect of taVNS on improving inhibitory control ability (Wright *et al*., 2014). Furthermore, the computational model assumes that the individual’s decision-making process after encoding the corresponding stimulus (such as sensory information encoding, etc.) begins from a starting point and accumulates information along the direction of two decision options until reaching the response boundary (Gomez *et al*., 2007; Ratcliff *et al*., 2018). The HDDM findings consistently showed that taVNS specifically decreased speed of information accumulation toward Go response (drift rate) in turn facilitating inhibitory control. These results suggest that taVNS may decelerate the top-down cognitive processing for Go responses during the present emotional Go/No-Go task, thereby breaking the pre-potent Go responses to improve response inhibition performance.

On the other hand, GLM results revealed increased activity in bilateral IFG, although stronger in the right side for angry expression faces, was found under active taVNS. It has been proposed that IFG, especially the right side, is a pivotal neural hub in regulating behavioral inhibition, and the activation of IFG was positively correlated with inhibitory control ability (Rubia *et al*., 2003; Aron *et al*., 2007; Chevrier *et al*., 2007; Zhuang *et al*., 2022). Considering that ADHD children show reduced activation in right IFG relative to normal controls during Go/No-Go task (Monden *et al*., 2012). taVNS may be a promising approach as treatment for ADHD. Additionally, activation of the right IFG is also associated with anger processing (Nomura *et al*., 2004; Taylor *et al*., 2018; Iarrobino *et al*., 2021; Sorella *et al*., 2021). By contrast, there were no significant changes under inhibition for happy expression faces following taVNS which may due to different pathways involved in inhibition of angry and happy stimuli. For example, differences in subcortical regions (i.e. striatum) activation have been found in positive and negative inhibition responses (Zhuang *et al*., 2021), although these regions cannot be detected using fNIRS. Thus overall, the enhancement of IFG activation suggests a critical role of IFG in modulating the effect of taVNS on emotional response inhibition, especially in the context of negative angry expression stimuli.

Notably, dynamic functional connectivity analyses also suggested that functional couplings (FCs) within prefrontal cortex including IFG, OFC, dlPFC and mPFC during the whole emotional inhibition task were significantly modulated by taVNS. This approach helps us quantify the experimental paradigm-based variations of brain functional networks affected by taVNS regardless of conditions (Tang *et al*., 2021; Lu *et al*., 2023). Among these couplings, taVNS significantly strengthened dlPFC-mPFC/ IFG/OFC/ dlPFC and OFC-IFG links in States1, 3 and 4, suggesting that such dlPFC- and OFC-related functional couplings played primary role in taVNS effects on emotional inhibitory processes. Most importantly, the positive impact of taVNS on inhibition performance was mediated by the increased functional coupling between IFG and OFC. Findings indicate that IFG-OFC coupling is involved in emotional inhibitory control. For example, individuals with higher strength IFG -OFC coupling were more flexible in regulating cognitive control and emotional processing (Shi *et al*., 2019) and the OFC plays a key role in serving inhibitory control in emotional context (Robertson *et al*., 2015; Stalnaker *et al*., 2015; Zhuang *et al*., 2021). Taken together, the increased FC between IFG and OFC might be the neural basis of enhanced inhibitory control for emotional faces under taVNS.

To further elucidate the neurocomputational mechanisms underlying the taVNS effect on emotional inhibitory control, a parallel mediation model was used and revealed that the positive effect of taVNS on inhibition performance was mediated by decreased drift rate for Go response in conjunction with increased FC between IFG and OFC. These results indicated that taVNS may improve response inhibition performance via its modulation of control ability for making Go responses more precise and enhancing FC within the prefrontal inhibitory control network.

Some limitations of the current study should be acknowledged. Firstly, some evidence from both animal and clinical studies proposed that taVNS would improve inhibitory control ability via modulating the LC-NE system (Burger *et al*., 2020; Colzato and Beste, 2020). Although prefrontal cortex circuits have been widely regarded as the center network of inhibition control ability (Goldstein *et al*., 2007) and LC-NE system could modulate the functional connectivity within the prefrontal inhibitory control network (Passamonti *et al*., 2018; Tomassini *et al*., 2022), given neural activities from subcortical regions cannot be detected through fNIRS technique, subcortical networks, such as basal ganglia, hypothalamus could be further investigated under taVNS, for example using deep brain stimulation techniques. Pupil diameter is also related to NE-LC system (Burger *et al*., 2020) although in our previous study we did not find any significant effects of taVNS on this (Zhu *et al*., 2022a). In addition, the current study only recruited healthy subjects and more clinical samples, especially individuals with deficits in inhibition control (e.g., ADHD, PTSD, addiction disorders) should be included to confirm and validate the positive effects of taVNS and their therapeutic potential.

In conclusion, the present study demonstrated a beneficial effect of taVNS on improving inhibitory control and further revealed the neurocomputational mechanisms underlying this effect in healthy individuals, suggesting a therapeutic potential of taVNS as a promising neuromodulation technique in the intervention of psychiatric disorders characterized inhibitory control deficits, such as attention deficit hyperactivity disorder, substance abuse disorder, and post-traumatic stress disorder.

## Data Availability

All data produced in the present study are available upon reasonable request to the authors.

## Acknowledgments

This work was supported by Natural Science Foundation of Sichuan Province [grant number 2022NSFSC1375 - WHZ], Fundamental Research Funds for the Central Universities, UESTC [grant number ZYGX2020J027 - WHZ], National Natural Science Foundation of China (NSFC) [grant number 31530032 - KMK], the Special Fund for Basic Scientific Research of Central Colleges (grant number ZYGX2021J036 - KMK) and Key Scientific and Technological projects of Guangdong Province [grant number 2018B030335001 - KMK].

## Availability of Data and Materials

Data in the present study can be made available upon request to the primary contact author, and code will be shared upon publication through a GitHub repository.

